# Evaluation of QuantiFERON SARS-CoV-2 interferon-γ release assay following SARS-CoV-2 infection and vaccination

**DOI:** 10.1101/2022.09.03.22279558

**Authors:** Síle A Johnson, Eloise Phillips, Sandra Adele, Stephanie Longet, Tom Malone, Chris Mason, Lizzie Stafford, Anni Jamsen, Siobhan Gardiner, Alexandra Deeks, Janice Neo, Emily J Blurton, Jemima White, Muhammed Ali, Barbara Kronsteiner-Dobramysl, Dónal T Skelly, Katie Jeffery, Christopher P Conlon, Philip Goulder, PITCH Consortium, Miles Carroll, Eleanor Barnes, Paul Klenerman, Susanna J Dunachie

**Affiliations:** Peter Medawar Building for Pathogen Research, Nuffield Department of Clinical Medicine, University of Oxford, Oxford, UK; University of Oxford Medical School, University of Oxford, Oxford, UK; University Hospital of Derby and Burton, UK; Oxford Centre For Global Health Research, Nuffield Department of Clinical Medicine, University of Oxford, Oxford, UK; Mahidol-Oxford Tropical Medicine Research Unit, Mahidol University, Bangkok, Thailand; Wellcome Centre for Human Genetics, Nuffield Department of Medicine, University of Oxford, Oxford, UK; Pandemic Sciences Institute, Nuffield Department of Medicine, University of Oxford, UK; Oxford University Hospitals NHS Foundation Trust, Oxford, UK; Department of Experimental Medicine, Nuffield Department of Clinical Medicine, University of Oxford, Oxford, UK; Nuffield Department of Clinical Neurosciences, University of Oxford, Oxford, UK; Radcliffe Department of Medicine, University of Oxford, Oxford, UK; Peter Medawar Building for Pathogen Research, Department of Paediatrics, University of Oxford, Oxford, UK; NIHR Oxford Biomedical Research Centre, University of Oxford, Oxford, UK; Translational Gastroenterology Unit, University of Oxford, Oxford, UK

**Keywords:** QuantiFERON, SARS-CoV-2, T cells, ELISpot, COVID-19

## Abstract

**Background:** T cells are important in preventing severe disease from SARS-CoV-2, but scalable and field-adaptable alternatives to expert T cell assays are needed. The interferon-gamma release assay QuantiFERON platform was developed to detect T cell responses to SARS-CoV-2 from whole blood with relatively basic equipment and flexibility of processing timelines.

**Methods:** 48 participants with different infection and vaccination backgrounds were recruited. Whole blood samples were analysed using the QuantiFERON SARS-CoV-2 assay in parallel with the well-established ‘Protective Immunity from T Cells in Healthcare workers’ (PITCH) ELISpot, which can evaluate spike-specific T cell responses.

**Aims:** The primary aims of this cross-sectional observational cohort study were to establish if the QuantiFERON SARS-Co-V-2 assay could discern differences between specified groups and to assess the sensitivity of the assay compared to the PITCH ELISpot.

**Findings:** The QuantiFERON SARS-CoV-2 distinguished acutely infected individuals (12-21 days post positive PCR) from naïve individuals (p< 0.0001) with 100% sensitivity and specificity for SARS-CoV-2 T cells, whilst the PITCH ELISpot had reduced sensitivity (62.5%) for the acute infection group. Sensitivity with QuantiFERON for previous infection was 12.5% (172-444 days post positive test) and was inferior to the PITCH ELISpot (75%).

Although the QuantiFERON assay could discern differences between unvaccinated and vaccinated individuals (55-166 days since second vaccination), the latter also had reduced sensitivity (55.5%) compared to the PITCH ELISpot (66.6%).

**Conclusion:** The QuantiFERON SARS-CoV-2 assay showed potential as a T cell evaluation tool soon after SARS-CoV-2 infection but has lower sensitivity for use in reliable evaluation of vaccination or more distant infection.

**Graphical abstract:** With the exception of acute infection group, the PITCH ELISpot S1+S2 had greater sensitivity for SARS-CoV-2 specific T cell responses compared with the QuantiFERON SARS-CoV-2 assay tube Ag3.

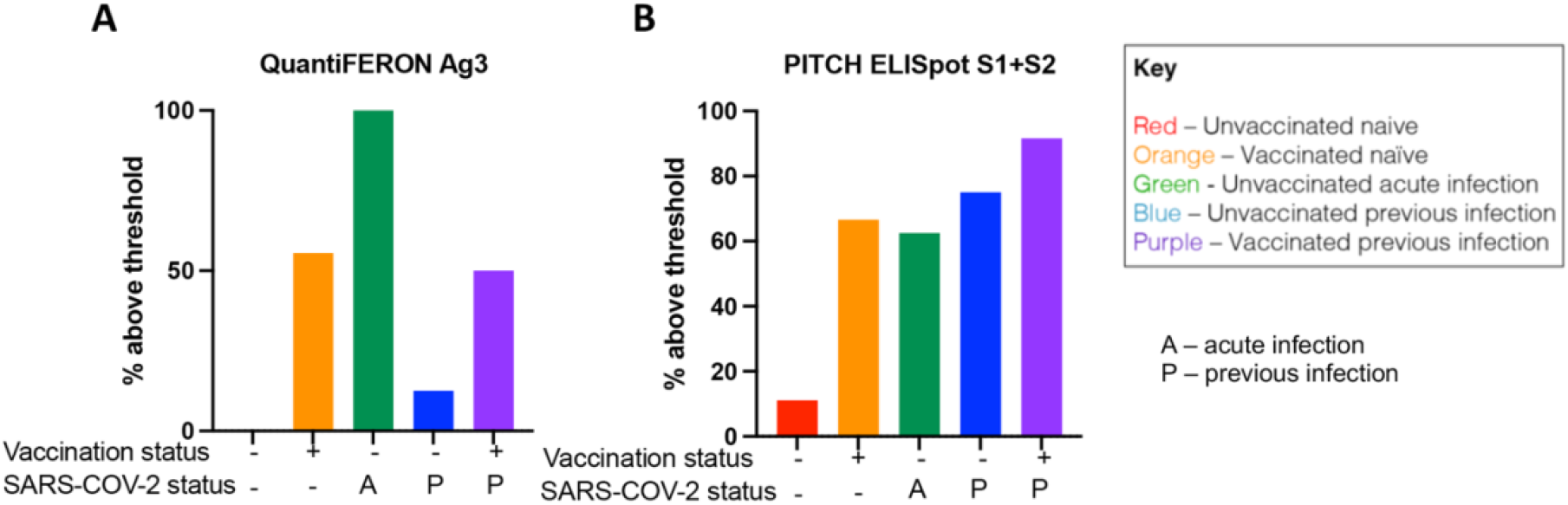

## Introduction

COVID-19 is a respiratory infection caused by SARS-CoV-2 with a recorded global burden of more than 500 million confirmed cases and over 6 million deaths (1). We and others have sought to characterise the immunological response to SARS-CoV-2 both following natural infection and vaccination (2–11). T cells are an important component of the immune response to SARS-CoV-2 infection and vaccination, persisting for several months post infection (4,8–10,12–16). T cells have also been extensively studied following vaccination alone (4,5,17–20) as well as in participants with combined past SARS-CoV-2 infection (5,17). Multiple assay platforms, including the *ex vivo* interferon-gamma enzyme-linked absorbent spot (IFN-γ ELISpot), Activation Induced Cell Marker (AIM), intracellular staining (ICS), T Cell proliferation assays and whole blood IFN-γ ELISA assays can be employed to evaluate T cell responses. Although these assays provide characterisation of T cell function, they can be time-consuming and require extensive laboratory reagents, equipment as well as expertise within hours of blood draw for reliable results.

The QuantiFERON SARS-CoV-2 assay is based on the well-characterised QuantiFERON TB interferon-gamma release assay (IGRA) (21). The basis for this platform is a whole blood cell-stimulation assay with plasma harvest for IFN-γ ELISA evaluation. The advantage of this platform is the workflow is straight forward to follow with tolerance for pause between steps which could accommodate various levels of expertise and diverse clinical/research settings including in low- and middle-income countries. A handful of studies have utilised the QuantiFERON SARS-CoV-2 assay (21–29). A small internal feasibility study carried out by Qiagen found quantifiable responses to vaccination over the course of four weeks after second vaccination (21). When analysing T cell responses after SARS-CoV-2 infection, the study showed detectable responses in three out of four participants with the assay. However, an important limitation of this small study was the lack of reference to a well-established cell-based assay to evaluate the potential of the QuantiFERON SARS-CoV-2 assay to accurately assess T cell responses.

Therefore, the present study sought to evaluate T cell responses using the QuantiFERON SARS-CoV-2 assay with parallel analysis using the well-established PITCH ELISpot (4–6,17,30) following SARS-CoV-2 infection and vaccination. The study also explored the sensitivity and specificity of the QuantiFERON SARS-CoV-2 assay in detecting SARS-CoV-2-specific T cell responses. Herein we present data to demonstrate a potential role for QuantiFERON SARS-CoV-2 as a reliable T cell evaluation tool soon after SARS-CoV-2 infection, but with low sensitivity compared to the conventional ELISpot assay for studying T cells following vaccination.

## Materials and Methods

### Study design and participant recruitment

Participants were sampled in the community or OPTIC (Oxford Protective T cell Immunity against COVID-19) study clinic in Oxford, UK once each between 9^th^ and 18^th^ June 2021. 48 participants were invited to participate by word of mouth and email communication of local healthcare workers, research scientists and students and informed consent was obtained under one of two studies: The GI Biobank Study 16/YH/0247, approved by the research ethics committee (REC) at Yorkshire & The Humber - Sheffield Research Ethics Committee on 29 July 2016, which had been amended for this purpose on 8 June 2020 or the Family Study R71346/RE001, approved by Oxford University’s Medical Sciences Inter-Divisional REC (MS-IDREC-R71346/RE00). Our target was 10 participants per group as a feasible number allowing meaningful statistical comparison, although no formal power calculation was performed.

Participants were sampled in the community or at the OPTIC (Oxford Protective T cell Immunity against COVID-19) study clinic in Oxford, UK once each during the month of June 2021. Following blood draw, samples for the QuantiFERON SARS-CoV-2 assay were kept at 4-8°C degrees for up to 48 hours before processing. The rest of the sample was used for isolation of peripheral blood mononuclear cells (PBMC) that were cryopreserved on the sample day and frozen for future use in the PITCH ELISpot assay. Participants were designated as naïve or previously infected for SARS-CoV-2 based on a positive PCR and/or serology at any time. Acute SARS-CoV-2 infection group was classified as blood sampling 12-21 days since a positive PCR test. For vaccination status, participants were designated as unvaccinated or vaccinated according to self-reported status. Serological status was determined using the Mesoscale Discovery (MSD) assay as described below, with a positive result for anti-S and/or anti-N supporting previous infection in an unvaccinated participant, and a positive result for anti-N supporting previous infection in a vaccinated participant (**Supplementary Figure 1**) meriting their exclusion from analysis.

### QuantiFERON SARS-CoV-2 assay

SARS-CoV-2-specific T cells were analysed using the QuantiFERON SARS-CoV-2 Research Use Only platform. The QuantiFERON SARS-CoV-2 Starter Pack (Qiagen, cat. no. 626115), Extended Pack (Qiagen, cat. no. 626215) and Control Set (Qiagen, cat. no. 626015) were employed, consisting of assay tubes coated with one of three sets of selected SARS-CoV-2 T cell antigens: Ag1 - CD4^+^ T cell epitopes from the S1 subunit (receptor binding domain) of the SARS-CoV-2 spike protein, Ag2 - CD4^+^ and CD8^+^ epitopes from the S1 and S2 subunits of the SARS-CoV-2 spike protein and Ag3 (Extended Pack) - CD4^+^ and CD8^+^ epitopes from S1 and S2, as in Ag2, but also immunodominant CD8^+^ epitopes of the whole proteome. The Control pack contains a ‘Nil tube’ which serves as the negative control and a ‘Mitogen tube’ which serves as a positive control.

The QuantiFERON SARS-CoV-2 kits were used in accordance with the manufacturer’s instructions. Whole blood samples, 0.8-1.2 ml, were collected directly into the assay collection tubes or into lithium heparin blood tubes for later transfer to the assay tubes. Assay tubes containing the whole blood were shaken and incubated for 16-24 hours at 37°C before centrifugation at 2,500 *g* for 15 minutes. Plasma was harvested from the top layer of the tube by gentle pipetting before being subjected to IFN-γ ELISA (Qiagen, cat. no. 626410). Following ELISA, quantitative results (IFN-γ concentration in IU/ml) were generated by subtracting the ‘Nil’ values from samples and interpolating values using an 8-parameter logistic model standard curve. In the absence of instruction from the manufacturer, the threshold to designate responses as positive was selected to be values greater than the mean of the Nil controls + 2 standard deviations, in keeping with a frequently used threshold for ELISpot assays (4,31). A total of 6-7 ml whole blood per participant time point was required for the three antigen tubes and controls. Samples were randomised for processing with the technician blinded to study group status to mitigate performance and verification bias.

### Isolation of peripheral blood mononuclear cells (PBMC), plasma and serum

PBMCs and plasma were isolated by density gradient centrifugation from 10 ml blood collected in EDTA tubes, and serum was collected in a serum-separating tube (SST, Becton Dickinson) as previously described (4). Briefly, PBMCs were isolated by density gradient centrifugation using Lymphoprep™ (p=1.077 g/ml, Stem Cell Technologies), washed twice with R0 (RPMI 1640 (Sigma, St. Louis, MO, USA) containing 10 mM Pen/Strep (100 U/mL) and 2 mM L-glutamine (100 µg/mL) (Sigma)) and resuspended in R10 (R0 supplemented with 10% FBS) or AutoMACs Rinse Buffer and counted using the Guava® ViaCount™ assay on the Muse Cell Analyzer (Luminex Cooperation). PBMCs were resuspended in freezing mix (FBS with 10% DMSO) and frozen down to −80°C before storage in liquid nitrogen. To obtain plasma, the uppermost fraction following the initial Lymphoprep centrifugation above was collected and centrifuged at 2000g for 10 minutes to remove platelets before storage at −80°C. Donor blood was also collected in a serum-separating tube (SST, Becton Dickinson) which was centrifuged at 2000g for 10 minutes. Serum was removed and stored at −80°C. As for the QuantiFERON testing, samples were randomised for processing with the technician blinded to study group status in order to mitigate performance and verification bias.

### In-house PITCH ELISpot assay

The PITCH ELISpot Standard Operating Procedure (SOP) is available as published previously (17). *Ex vivo* IFN-γ ELISpot assays were set up from cryopreserved peripheral blood mononuclear cells (PBMCs) using the Human IFN-γ ELISpot Basic kit (Mabtech 3420-2A). MultiScreen-IP filter plates (Millipore, MAIPS4510) were coated with 50 µl/well using the ELISpot Basic Kit Capture antibody (clone 1-D1K) at 10 μg/ml diluted in sterile phosphate buffered saline (PBS; Fisher Scientific) or sterile carbonate bicarbonate (Sigma Aldrich) for 8 to 48 h at 4°C. PBMCs were thawed and resuspended in Rab10 (filtered R0 media (Sigma) supplemented with 10% Human serum) with DNase, and allowed to rest for 2-3 h in an incubator at 37°C, 5% CO_2_, 95% humidity prior to stimulation with peptides. The capture antibody coated plates were washed twice with R0, then blocked with 100 µL/well of Rab10 for 1/2-8 h at RT or 8-48h at 4°C. Rested cells were centrifuged and resuspended in 1 ml Rab10 for counting on Muse™ Cell Analyser or Bio-Rad TC10TM Automated Cell Counter. After blocking, overlapping peptide pools (18-mers with 10 amino acid overlap Mimotopes) representing the spike (S1+S2), Membrane (M) or nucleocapsid (NP) SARS-CoV-2 proteins were added to 200,000 PBMCs/well at a final concentration of 2 μg/ml for 16 to 18 h. S1 and S2 were added in separate test wells, M and NP were combined in a singular test well. Pools consisting of CMV, EBV and influenza peptides at a final concentration of 2 μg/ml (CEF; Proimmune) and concanavalin A (ConA) at a final concentration of 5 μg/ml were used as positive controls. DMSO was used as the negative control at the equivalent concentration to the peptides. After cell stimulation overnight, wells were washed 7 times 100-200 µL/well with PBS with 0.05% (v/v) Tween20 (Sigma Aldrich) and incubated with 50 µL/well of the ELISpot Basic kit biotinylated detection antibody (clone 7-B6-1) diluted in PBS at 1 μg/ml, for 2-4 h at room temperature (RT). Wells were then washed 7 times with 100-200uL/well PBS-0.05% (v/v) Tween20, and then incubated with 50 µL/well of the ELISpot Basic kit streptavidin-ALP, diluted in PBS at 1 μg/ml for 1-2 h at RT. Wells were then washed 7 times with 100-200 µL/well PBS-0.05% Tween20 and colour development was carried out using the 1-step NBT/BCIP Substrate Solution. 50 µl of filtered NBT/BCIP was added to each well for 5-7 minutes in the dark at RT. Colour development was stopped by washing the wells with cold tap water. Air dried plates were scanned and analysed with the CTL Cellular Technologies Series 6 ALFA. Antigen-specific responses were quantified by subtracting the mean spots of the control wells from the test wells and the results were expressed as spot-forming units (SFU)/10^6^ PBMCs. Responses were defined as positive if values were greater than the mean of the DMSO control + 2 standard deviations with a minimum of 20 SFCs per 1 million PBMCs (4,31).

### Mesoscale Discovery (MSD) binding assay

IgG responses to SARS-CoV-2 were measured using a multiplexed MSD immunoassay: The V-PLEX COVID-19 Coronavirus Panel 3 (IgG) Kit from Meso Scale Diagnostics, Rockville, MD USA. A MULTI-SPOT® 96-well, 10 spot plate was coated with three SARS CoV-2 antigens (S, RBD, N) and bovine serum albumin. Antigens were spotted at 200−400 μg/mL (MSD® Coronavirus Plate 3). Multiplex MSD assays were performed as per the instructions of the manufacturer. To measure IgG antibodies, 96-well plates were blocked with MSD Blocker A for 30 minutes. Following washing with washing buffer, samples diluted 1:1,000-10,000 in diluent buffer, or MSD standard or undiluted internal MSD controls, were added to the wells. After 2 h incubation and a washing step, detection antibody (MSD SULFO-TAG Anti-Human IgG Antibody, 1/200) was added. Following washing, MSD GOLD Read Buffer B was added and plates were read using a MESO® SECTOR S 600 Reader. The standard curve was established by fitting the signals from the standard using a 4-parameter logistic model. Concentrations of samples were determined from the electrochemiluminescence signals by back-fitting to the standard curve and multiplied by the dilution factor. Concentrations are expressed in Arbitrary Units/ml (AU/ml). Cut-offs were determined for each SARS-CoV-2 antigen (S, RBD and N) based on the concentrations measured in 103 pre-pandemic sera + 3 Standard Deviation as previously published (5). Cut-off for S: 1160 AU/ml; cut-off for RBD: 1169 AU/ml; cut-off for N: 3874 AU/ml. Samples were processed blindly to mitigate performance and verification bias.

### Statistical analyses

Data were analysed by non-parametric tests: Mann-Whitney for non-paired comparisons between two groups and Kruskal Wallis with Dunn’s correction for comparisons between multiple groups. Correlation studies to compare values from different assays was calculated using Spearman correlation coefficient. To calculate sensitivity, specificity, positive predictive value and negative predictive value, the gold standard was designated as the ‘clinical phenotype’(CP) – self-reported vaccination or infection, with input from the MSD antibody binding assay. The threshold above which a sample was designated as ‘positive’ was designated as mean of the negative + 2 SD (4,31). Five tests were assessed: QuantiFERON SARS-CoV-2 Ag1, Ag2 and Ag3 and PITCH ELISpot S1+S2 and M+NP. For any of the given tests, a true positive (TP) was designated as being above threshold in CP+. A true negative (TN) was below threshold in CP−. A false positive (FP) was above threshold in CP−. A false negative was below threshold in CP+. To calculate sensitivity, specificity, positive predictive values (PPV) and negative predictive values, a 2×2 table was designed for each test and the following formulae applied: sensitivity = TP/TP+FN; specificity = TN/TN+FP; PPV = TP/TP+FP and NPV = TN/TN+FN.

GraphPad Prism v9.1.0 was used for statistical analysis and graphical representation.

## Results

### Participants of the study

Participants were recruited in June 2021 when the delta variant of SARS-CoV-2 was the dominant variant (32). Participants were assigned into 5 groups based on previous SARS-CoV-2 infection and vaccination status: Unvaccinated Naïve, Vaccinated Naïve, Unvaccinated Acute Infection, Unvaccinated Previous Infection and Vaccinated Previous Infection. Prior to data analysis, anti-S and anti-N antibodies were measured by Meso Scale discovery (MSD) assay to exclude asymptomatic previous infection status (**Supplementary Figure 1**) resulting in two participants being excluded from further analysis. One was excluded from the ‘unvaccinated naïve’ as they had positive Spike IgG, suggesting previous infection/vaccination. The other was excluded from the ‘vaccinated naïve’ group as they had positive N IgG suggesting previous infection. None of the participants who had previous infection required hospitalisation. Demographic information about the 46 participants included in the analysis is shown in **Table 1**. The age of the participants ranged from 18 to 56 with a median age of 24. Unvaccinated individuals were younger than vaccinated individuals (median 23 v median 28 respectively, p = 0.002) due to the progress of the national vaccination programme at the time of sampling (**Table 1**). There was no statistically significant difference between the median age of naïve and previously infected (27 v. 24 respectively, p = 0.18). In general, gender balance was achieved – except for the unvaccinated, acute infection group who were all participants from an outbreak in a local women’s rugby team. The participants in all groups predominantly identified as being of ‘white’ ethnicity. Most recipients received the Pfizer/BioNTech BNT162b2 vaccine reflecting the national vaccination roll out in the UK for this age group, with the remainder receiving the Oxford/AstraZeneca AZD1222 vaccine.

**Table 1.** Characteristics of participants in this study

|  | Unvaccinated<br>Naive | Vaccinated<br>Naive | Unvaccinated<br>Acute infection | Unvaccinated<br>Previous<br>infection | Vaccinated<br>Previous<br>infection |
| --- | --- | --- | --- | --- | --- |
| **<u>Number of subjects</u>** | **9** | **9** | **8** | **8** | **12** |
| **<u>Age, median (range)</u>** | **23 (22, 37)** | **30.1 (27, 46)** | **26 (19, 33)** | **23 (18, 24)** | **25.2 (21, 56)** |
| **<u>Gender</u>** |  |  |  |  |  |
| Male | 5 (55.5%) | 3 (33.3%) | 0 (0%) | 4 (50%) | 6 (50%) |
| Female | 4 (44.4%) | 6 (66.6%) | 8 (100%) | 4 (50%) | 6 (50%) |
| Other | 0 (0%) | 0 (0%) | 0 (0%) | 0 (0%) | 0 (0%) |
| **<u>Ethnicity</u>** |  |  |  |  |  |
| White | 5 (55.5%) | 7 (77.7%) | 7 (87.5%) | 7 (87.5%) | 9 (75%) |
| Black | 1 (11.1%) | 0 (0%) | 0 (0%) | 0 (0%) | 1 (8.33%) |
| Asian | 2 (22.2%) | 1 (11.1%) | 0 (0%) | 1 (12.5%) | 1 (8.33%) |
| Mixed | 1 (11.1%) | 0 (0%) | 1 (12.5%) | 0 (0%) | 1 (8.33%) |
| Other | 0 (0%) | 1 (11.1%) | 0 (0%) | 0 (0%) | 0 (0%) |
| **<u>Vaccine</u>** |  |  |  |  |  |
| Pfizer BNT162b2 | N/A | 8 (88.8%) | N/A | N/A | 8 (75%) |
| Oxford/AstraZeneca<br>AZD1222 | N/A | 1 (11.1%) | N/A | N/A | 4 (25%) |
| **<u>Days since V2, median</u>**<br>**<u>(range)</u>** | **N/A** | **102 (55, 166)** | **N/A** | **N/A** | **65 (54, 160)** |
| **<u>Days since SARS-COV-2,</u>**<br>**<u>median (range)</u>** | **N/A** | **N/A** | **16 (12, 21)** | **256 (172, 444)** | **221 (145,**<br>**433)** |

### QuantiFERON SARS-CoV-2 in SARS-CoV-2 infected individuals

To examine the QuantiFERON SARS-CoV-2 assay in SARS-CoV-2-infected individuals, samples from the unvaccinated naïve group were compared to unvaccinated, acute infection samples (median of 16 days, range 12-21 days since positive SARS-CoV-2 PCR). The same unvaccinated naïve samples were also compared to unvaccinated, previous infection samples (median of 256, range 172-444 days since positive test). For all three QuantiFERON assay tubes, Ag1, Ag2 and Ag3, there was significantly greater IFN-γ detected for acute infection individuals compared to naïve controls (**Figure 1A**; p < 0.0001 for all). Samples were also compared using two PITCH ELISpot assays – one against the spike protein (S1+S2) and one against structural proteins M protein and nucleocapsid protein (M+NP). A significant difference was also seen in the PITCH ELISpot comparison for S1+S2 and M+NP between naïve and acute infection groups (**Figure 1B**; p = 0.037 and p = 0.019 respectively). For the naïve vs previous infection group comparison, there was no statistically significant difference in the amount of IFN-γ produced in any of the three QuantiFERON tubes (**Figure 1C**), although the PITCH ELISpot was able to detect differences in S1+S2 (p = 0.029) and M+NP (p = 0.007) when comparing the two groups (**Figure 1D**). When looking at vaccinated individuals and aiming to differentiate SARS-CoV-2 infection naïve and previous infection (median of 222 days, range 175-433 days since positive test), there were no statistically significant differences between the groups using QuantiFERON Ag1, Ag2 or Ag3 (**Figure 1E**). The PITCH ELISpot found a difference between the vaccinated naïve and vaccinated previous infection groups with M+NP (p = 0.0005) but not for S1+S2 (p = 0.254) (**Figure 1F**).

**Figure 1.**
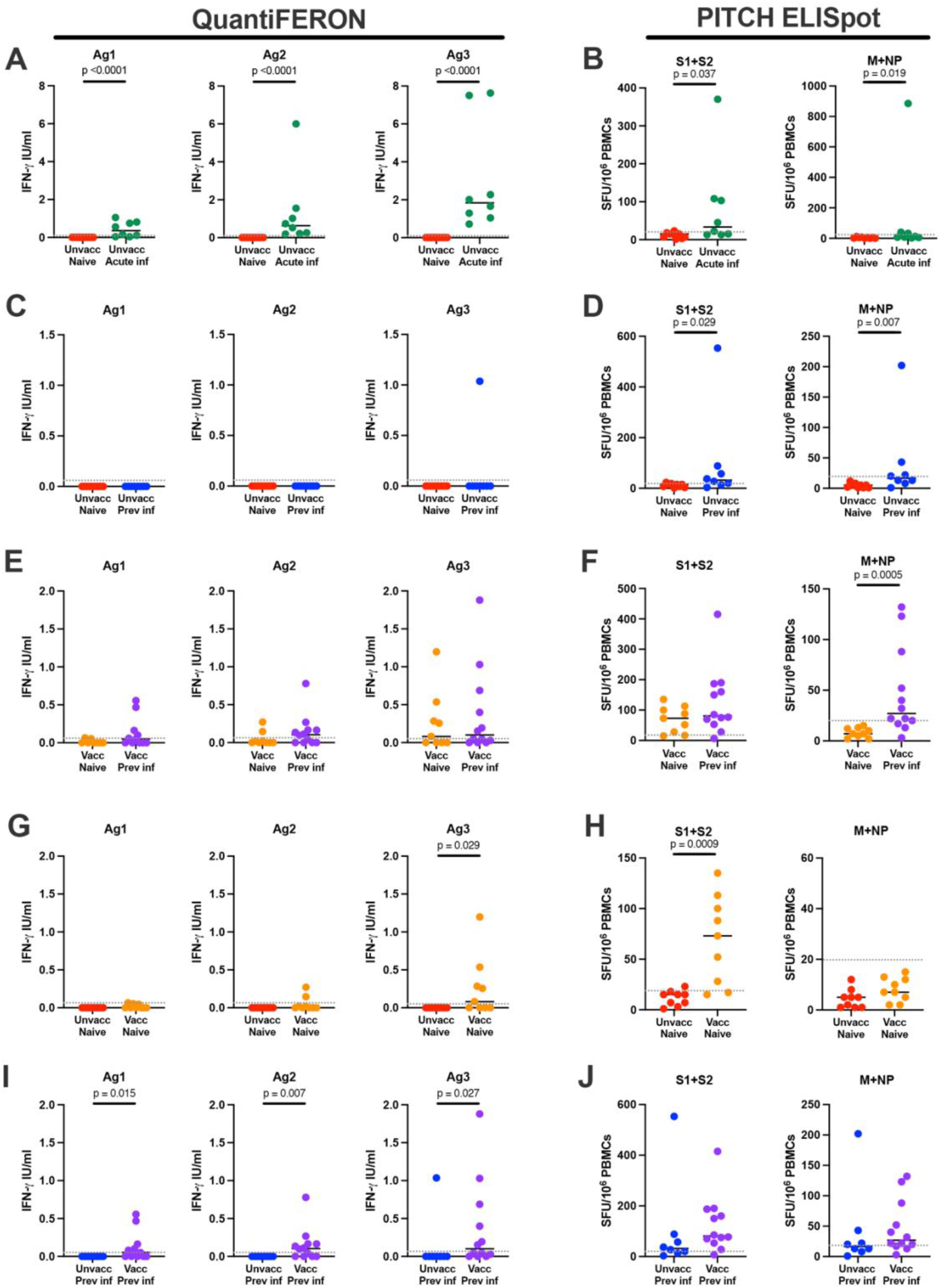
Comparison of T cell responses between indicated groups measured by QuantiFERON and PITCH ELISpot. (A) T cell responses to SARS-CoV-2 in unvaccinated naïve and unvaccinated acute infection using QuantiFERON Ag1, Ag2 and Ag3, and (B) ELISpot S1+S2 and M+NP. (C) T cell responses to SARS-CoV-2 in unvaccinated naïve and unvaccinated previous infection using QuantiFERON Ag1, Ag2 and Ag3, and (D) ELISpot S1+S2 and M+NP. (E) T cell responses to SARS-CoV-2 in vaccinated naïve and vaccinated previous infection using QuantiFERON Ag1, Ag2 and Ag3, and (F) ELISpot S1+S2 and M+NP. (G) T cell responses to SARS-CoV-2 in unvaccinated naïve and vaccinated naive using QuantiFERON Ag1, Ag2 and Ag3, and (H) ELISpot S1+S2 and M+NP. (I) T cell responses to SARS-CoV-2 in unvaccinated previous infection and vaccinated previous infection using QuantiFERON Ag1, Ag2 and Ag3, and (J) ELISpot S1+S2 and M+NP. Unvacc naïve – unvaccinated naïve, n = 9; unvacc acute inf - unvaccinated acute infection, n = 8; unvacc prev inf - unvaccinated previous infection, n = 8; vacc naïve - vaccinated naïve, n = 10; vacc prev inf – vaccinated previous infection, n = 12. Unpaired comparisons between groups were performed using Mann-Whitney test, with statistical significance as p <0.05. Horizontal dotted lines represent the threshold of each assay based on negative assay controls (see Methods).

### QuantiFERON SARS-CoV-2 in individuals vaccinated against SARS-CoV-2

To examine the QuantiFERON SARS-CoV-2 assay in vaccinated individuals, we compared samples from unvaccinated naïve versus vaccinated naïve individuals (median of 102, range 55-166 days since second vaccination). Here, there was no difference between the two groups for Ag1 (p = 0.206) or Ag2 (p = 0.082) but there was a statistically significant difference for Ag3 (p = 0.029) (**Figure 1G**). The PITCH ELISpot demonstrated differences between the groups for S1+S2 (p = 0.0005) but not M+NP (p = 0.072) (**Figure 1H**).

We also compared SARS-CoV-2 previously infected individuals with and without vaccination, (median of 65, range 54-160 days since second vaccination). The QuantiFERON assay was able to detect statistically significant differences between the two groups for all three assay tubes; Ag1 (p = 0.015), Ag2 (p = 0.007) and Ag3 (p = 0.027) (**Figure 1I**) – largely due to a lack of responses in the previously infected unvaccinated cohort. The PITCH ELISpot did not present differences between the groups for either S1+S2 (p = 0.115) or M+NP (p = 0.245) (**Figure 1J**).

### Sensitivity of the QuantiFERON SARS-CoV-2 assay

The QuantiFERON SARS-CoV-2 assay utilises the QuantiFERON IGRA technology, known for its use in detecting tuberculosis with QuantiFERON TB Gold. This assay has a standardised threshold for designating samples as being ‘positive’. As such, we sought to present the current data as qualitative positive or negative results. As there was no threshold provided with the QuantiFERON SARS-CoV-2 assay, we utilised the threshold established for ELISpot assays: the mean of the negative control samples (in this case, the ‘Nil’ tubes) plus two standard deviations, as previously published (4,31). Samples above threshold were used to determine the sensitivity of a given test for SARS-CoV-2 T cell responses.

Using this threshold, for Ag1, none of the unvaccinated naïve samples were above threshold (**Figure 2A**), with 22.2% of vaccinated naïve, 75% of unvaccinated acute infection, none of unvaccinated previous infection, and 58.3% of vaccinated previous infection above threshold. For Ag2, the results were similar to Ag1: none of the naïve samples were above threshold, with 22.2% of vaccinated naïve, 100% of unvaccinated acute infection, none of unvaccinated previous infection and 58.3% of vaccinated previous infection above threshold (**Figure 2B**). For Ag3, none of the unvaccinated naïve samples were above threshold. 55.5% of vaccinated naïve samples were above threshold, 100% unvaccinated acute infection, 12.5% of unvaccinated previous infection and 50% of vaccinated previous infection were above threshold (**Figure 2C**). The S1+S2 PITCH ELISpot showed greater sensitivity for SARS-CoV-2 T cell responses in general (**Figure 2D**). 11.11% of unvaccinated naïve samples were above threshold, with 66.6% of vaccinated naïve, 62.5% of unvaccinated acute infection, 75% of unvaccinated previous infection and 91.67% of vaccinated previous infection above threshold. For M+NP, there was less sensitivity compared to S1+S2 with 0% of naive unvaccinated and naive vaccinated above threshold (**Figure 2E**), 10% of vaccinated naïve, 37.5% of unvaccinated acute infection and unvaccinated previous infection, and 91.67% of vaccinated previous infection above threshold.

**Figure 2.**
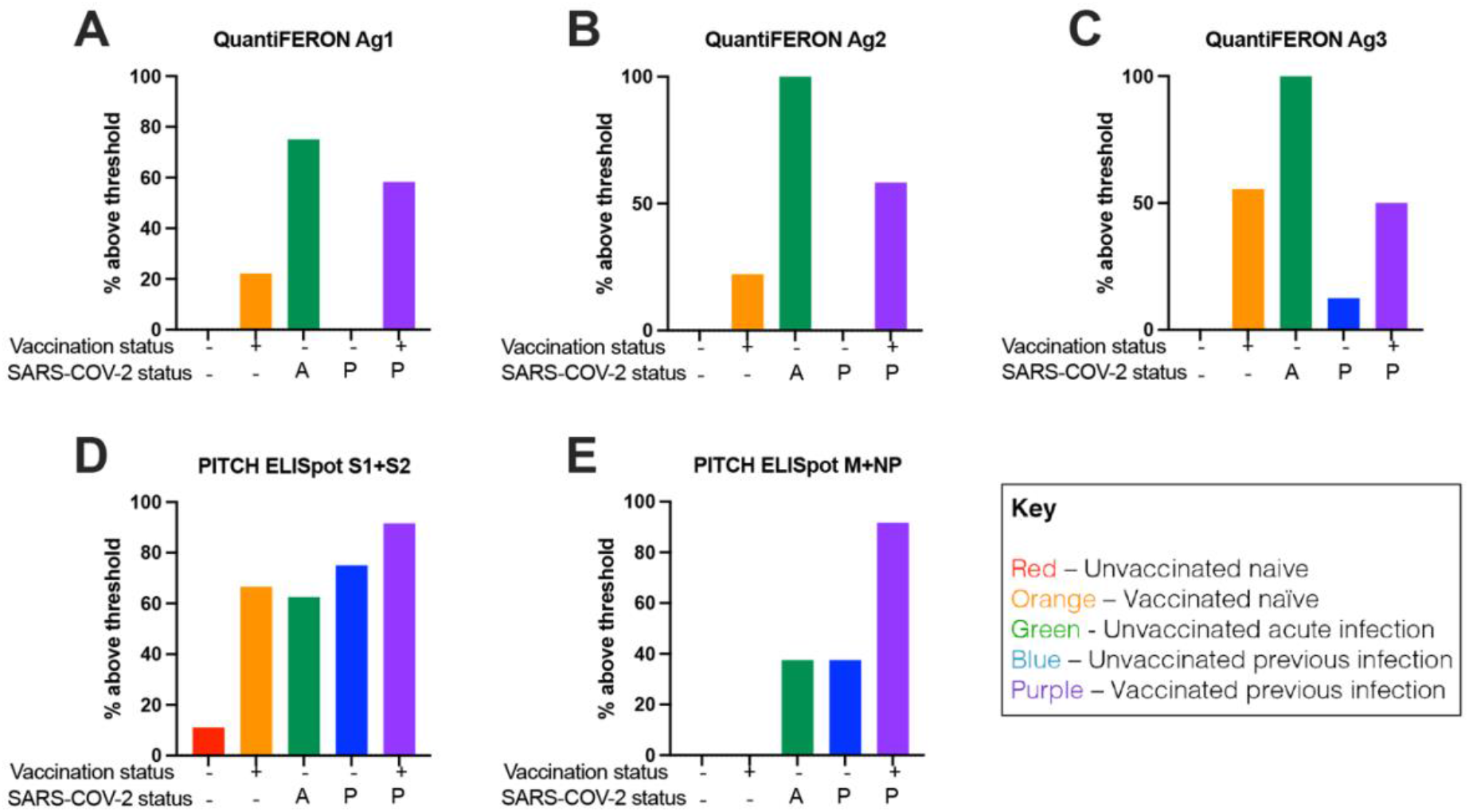
Percentage of samples above threshold for indicated groups using QuantiFERON and PITCH ELISpot. The threshold to designate responses as positive was selected to be values greater than the mean of the Nil controls + 2 standard deviations, with a minimum of 20 SFCs per million PBMCs for ELISpot. Percentage of samples above threshold for all five groups using (A) QuantiFERON Ag1 (B) QuantiFERON Ag2 (C) QuantiFERON Ag3 (D) PITCH ELISpot S1+S2 (E) PITCH ELISpot M+NP. Unvaccinated naïve, n = 9; unvaccinated acute infection, n = 8; unvaccinated previous infection, n = 8; vaccinated naïve, n = 10; vaccinated previous infection, n = 12. A – acute infection; P – previous infection.

Sensitivity, specificity, positive predictive values and negative predictive values of all groups for each of the five tests are detailed in **Table 2** with clinical positive (CP) phenotypes determined on the basis of historic infection and/or vaccination, with the MSD antibody assay removing those reported naïve which had positive antibody responses. For unvaccinated naïve samples, all three QuantiFERON SARS-CoV-2 assay tubes had 100% specificity and 100% negative predictive value (NPV) for SARS-CoV-2 T cells, as did the PITCH ELISpot M+NP but PITCH ELISpot S1+S2 had only 88.9% specificity. For vaccinated naïve samples, PITCH ELISpot S1+S2 had greater sensitivity (66.6%) than any of the QuantiFERON tubes Ag1, Ag2 or Ag3 (22.2%, 22.2% and 55.5% respectively). For this group, all tests had 100% positive predictive value for SARS-CoV-2 T cells. For the unvaccinated acute infection group, all five tests exhibited 100% PPV, and as stated before, the QuantiFERON SARS-CoV-2 Ag2 and Ag3 exhibited 100% sensitivity for SARS-CoV-2 T cells whist Ag1 had 75% and PITCH ELISpot S1+S2 and M+NP had 62.5% and 37.5% respectively. For the unvaccinated previous infection group, as before the PITCH ELISpot had superior sensitivity with S1+S2 at 75% and M+NP at 37.5% whilst the QuantiFERON SARS-CoV-2 achieved 0%, 0% and 12.5% for Ag1, Ag2 and Ag3 respectively. Finally, there was greater sensitivity for SARS-CoV-2 T cells using the PITCH ELISpot (91.67% for both S1+S2 and M+NP) than QuantiFERON SARS-CoV-2 which only achieved 58.33% sensitivity for Ag1 and Ag2, and only 50% for Ag3. All tests for this group had 100% PPV.

**Table 2.**
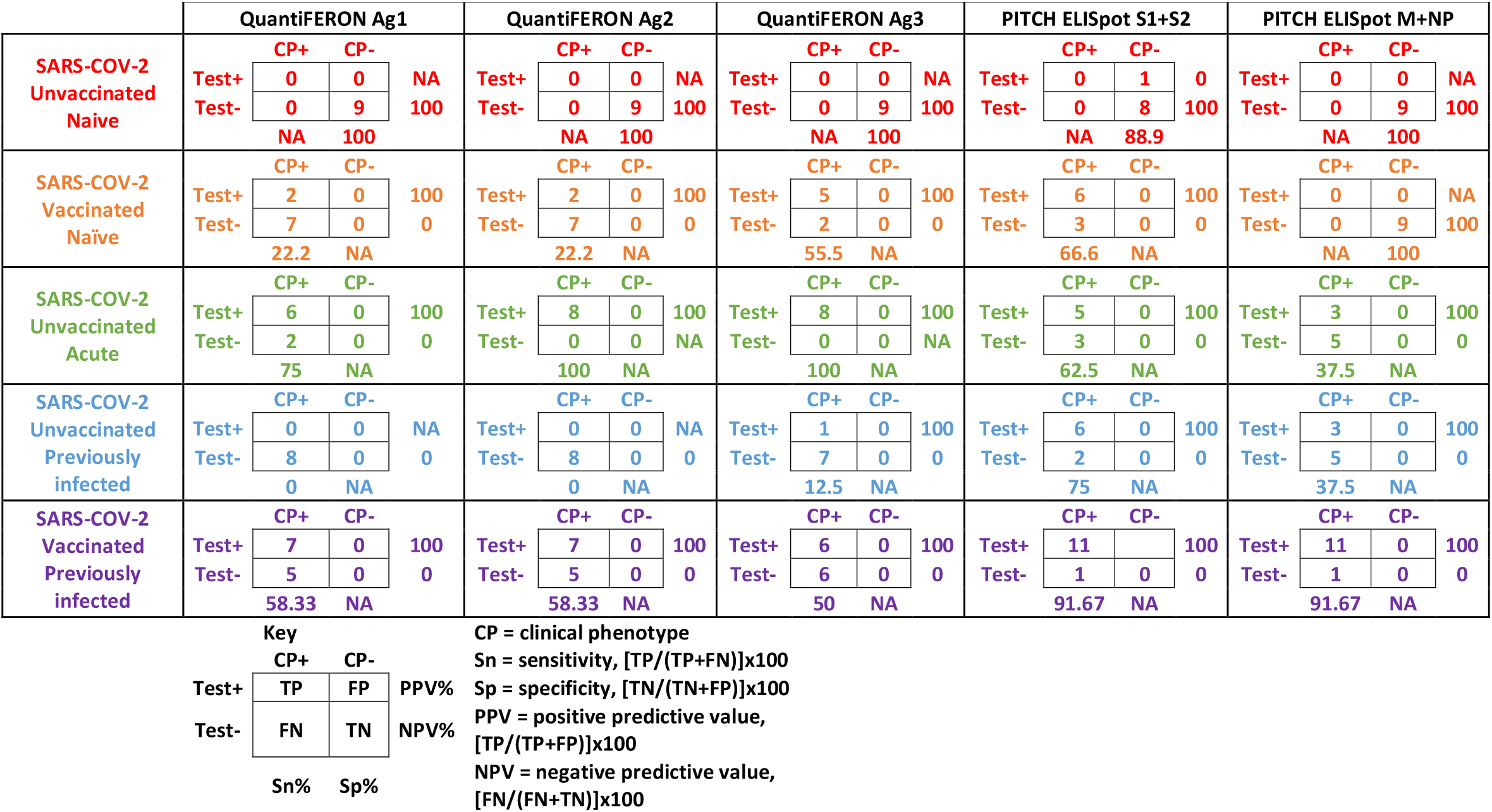
Sensitivity, specificity, positive predictive values and negative predictive vales for QuantiFERON and ELISpot assays. TP, true positive; FP, false positive; TN, true negative; FN, false negative.

### Correlation of QuantiFERON SARS-CoV-2 assay tubes

Correlation analysis was performed between the three assay tubes for the QuantiFERON SARS-CoV-2 (**Supplementary Figure 2**). For all three comparisons (Ag1 v Ag2, Ag1 v Ag3 and Ag2 v Ag3) there was significant correlation (R^2^ 0.7132 – 0.889, p < 0.0001).

### Correlation of the QuantiFERON SARS-CoV-2 assay with PITCH ELISpot and MSD antibody platform

Correlation analysis was performed between the three QuantiFERON SARS-CoV-2 assay tubes and the PITCH ELISpot assay. For all three assay tubes there was no significant correlation with PITCH ELISpot S1+S2 (**Figure 3A-C**) or with PITCH M+NP (**Figure 3D-F**). The QuantiFERON SARS-CoV-2 Ag1, Ag2 and Ag3 showed statistically significant correlation with S IgG (**Figure 4A-C**) RBD IgG (**Figure 4D-F**) and N IgG (**Figure 4G-I**).

**Figure 3.**
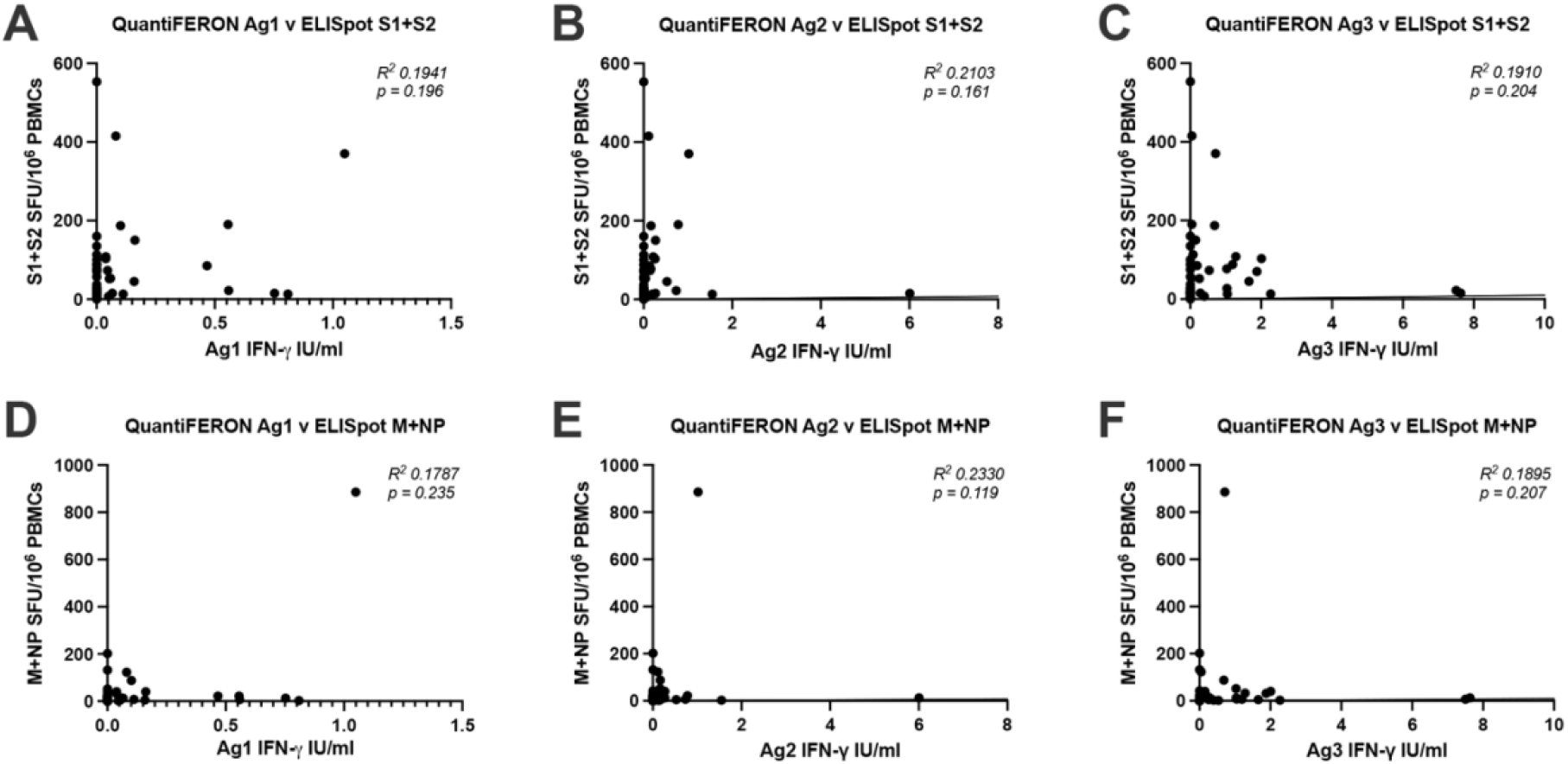
Correlation between T cell responses measured by QuantiFERON and PITCH ELISpot. (A-C) Correlation between QuantiFERON Ag1, Ag2 and Ag3 with ELISpot S1+S2. (D-F) Correlation between QuantiFERON Ag1, Ag2 and Ag3 with ELISpot M+NP. N = 47 for each analysis. Correlation analysis was carried out with Spearman’s r correlation and two-tailed P values reported, with alpha = 0.05.

**Figure 4.**
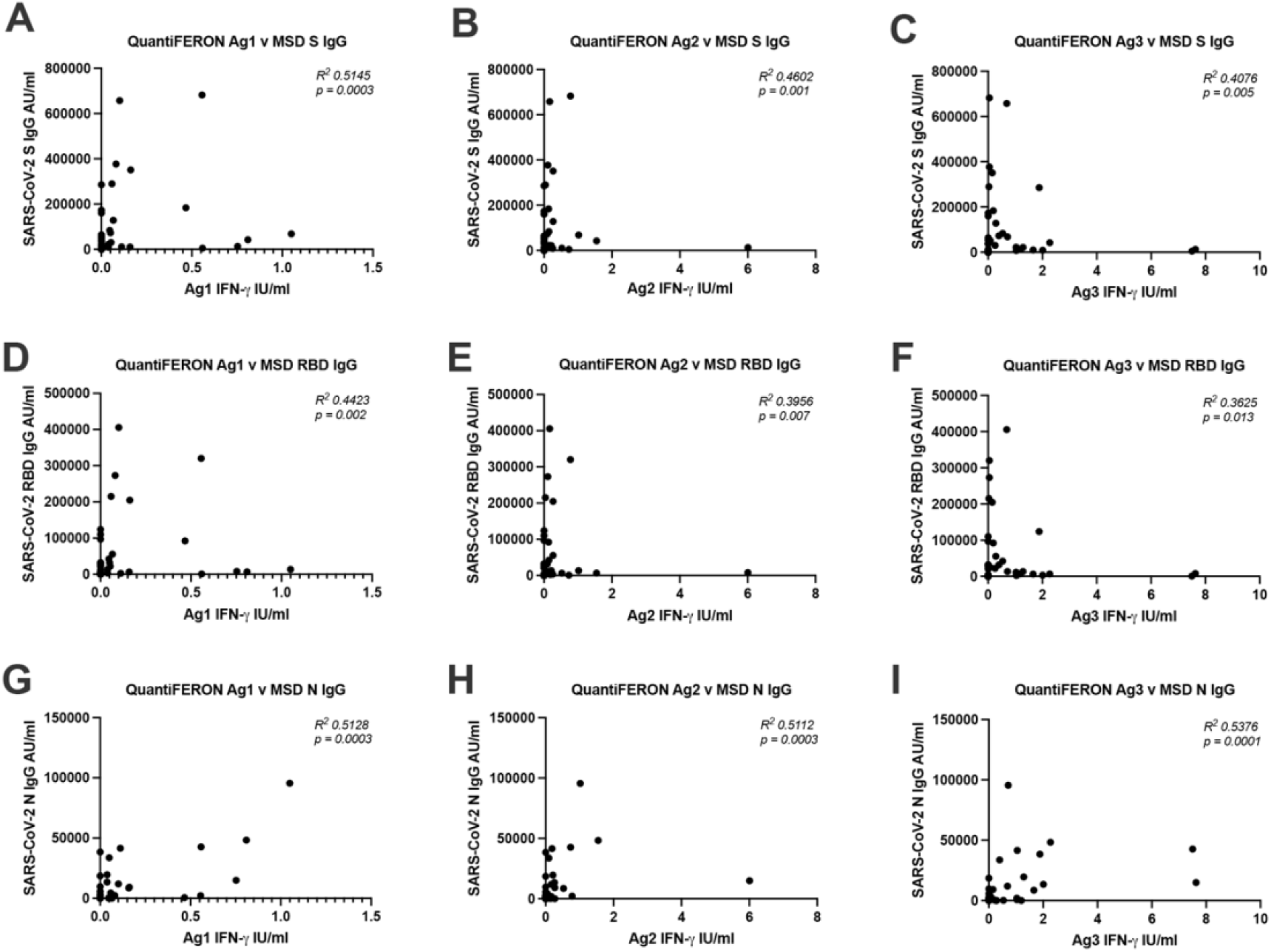
Correlation between T cell responses measured by QuantiFERON and antibodies measured by MSD binding assay. (A-C) Correlation between QuantiFERON Ag1, Ag2 and Ag3 with MSD S IgG. (D-F) Correlation between QuantiFERON Ag1, Ag2 and Ag3 with MSD RBD IgG. (G-I) Correlation between QuantiFERON Ag1, Ag2 and Ag3 with MSD N IgG. N = 47 for each analysis. Correlation analysis was carried out with Spearman’s r correlation and two-tailed P values reported, with alpha = 0.05.

## Discussion

T cells are increasingly recognised for their role in SARS-CoV-2 infection and vaccination (33–36). However, cell-based assays which evaluate T cells are typically labour- and expertise-intensive, require specialist equipment, and need specialist processing of fresh blood within 3-4 hours of blood draw. Therefore, validating simple, commercially available kits could expand the repertoire of tools for evaluating T cells in the context of SARS-CoV-2, particularly in research laboratories which do not have means to overcome the above barriers. The present study sought to do this with the commercially available QuantiFERON SARS-CoV-2 assay, which is based on the same technology of the QuantiFERON TB Gold used worldwide (reviewed in (37)). This assay has a straight-forward work-flow, basic equipment requirements compared to other cell-based assays and tolerance of delays in processing. Moreover, the read-out can be seen at times with the naked eye, which may merit further investigation (**Supplementary Figure 4**).

The data presented in this study demonstrates a robust read-out for all three QuantiFERON assay tubes (Ag1, Ag2, Ag3) for acute infection samples within 21 days of positive PCR, with superior sensitivity for SARS-CoV-2 T cell responses than both the S1+S2 and M+NP ELISpot for these samples. Furthermore, none of the naïve samples generated a positive result, making the QuantiFERON SARS-CoV-2 highly specific. These results support a utility for the QuantiFERON SARS-CoV-2 in evaluating T cell responses during or soon after acute infection. The QuantiFERON SARS-CoV-2 was unable to detect robust responses in individuals who had tested positive for SARS-CoV-2 infection 172-444 days earlier, suggesting the possibility of a restricted time frame for detection using this assay. Effector T cell responses to SARS-CoV-2 infection decrease over time (8,13) which may explain why T cell responses were not detected in more distant infection, but are typically still detectable by research assays more than 6 months later (20,38,39). Further work to increase the sensitivity of the QuantiFERON SARS-CoV-2 assay to detect SARS-CoV-2 T cell responses in this group would be useful for evaluating T cell responses in distant infection.

The highly sensitive and specific results in the acute infection samples show potential for the assay to be used as a diagnostic test, as is the case for the QuantiFERON TB IGRA in settings where PCR testing may not be feasible. T cells can be detected as soon as 3-5 post symptom onset, with a similar kinetic profile to antibody detection (40,41), which supports the use of T cell-based diagnostic SARS-CoV-2 tests. However, the utility of such a diagnostic test would be at the time of symptom onset, which was not possible to assess in the current study due to national isolation guidelines at the time of sampling. Further studies closer to symptom onset are required to investigate further.

The low responses to Ag1, Ag2 or Ag3 seen in naïve participants post vaccination limit the utility of the QuantiFERON assay for scaled up study of response to vaccination. This is disappointing when there is a huge need for large scale prospective longitudinal studies in a range of populations and settings, to establish immune correlates of protection and differences in vaccine response between vulnerable patient groups. A larger dynamic range of IFN-γ responses post vaccination or infection has been observed in another whole blood ELISA-based assay (9,10), although some of these samplings were taken closer to the time of vaccination which may explain their greater sensitivity for SARS-CoV-2 T cell responses than in the present study. Further work to raise the sensitivity of the QuantiFERON assay, such as increasing the detection of IFN-γ by ELISA, would be hugely valuable and would have high potential for transfer to other emerging outbreak pathogens in regions of the world with limited laboratory capacity. Nevertheless, the presence or absence of a T cell response detectable by the QuantiFERON assay could prove to be a useful parameter to include in longitudinal studies of vaccine immunogenicity and correlates of protection.

With the exception of the acutely infected group, the PITCH ELISpot S1+S2 exhibited superior sensitivity for SARS-CoV-2 T cell responses compared to the QuantiFERON SARS-CoV-2 assay in keeping with other studies which have demonstrated the value of ELISpot compared to other T cell evaluation tools (42,43). Likely factors contributing to the relatively inferior performance of the QuantiFERON platform could include the T cell concentration in whole blood samples, processing timelines and concentrations/selection of epitopes for T cell stimulation. However, much of the information required to draw strong conclusions is proprietary information, thus limiting our understanding for the performance differences between the platforms.

According to the manufacturer, the epitopes lining the Ag1 assay tube activate CD4^+^ T cells specific to RBD, Ag2 activates CD4^+^ and CD8^+^ T cells specific to S1 and S2, and Ag3 activates CD4^+^ and CD8^+^ T cells against numerous SARS-CoV-2 peptides including spike. In the five sets of two-group comparisons illustrated in **Figure 1**, Ag1 was able to discern statistically significant differences in two of the five group comparisons, Ag2 in three of five and Ag3 in three of five. In terms of sensitivity, Ag3 had the highest sensitivity for SARS-CoV-2-specific T cells in four of the five individual groups. Overall, there was correlation between the QuantiFERON SARS-CoV-2 assay tubes. Taken together, the present data suggests the Ag3 tube may be the most useful of the three for evaluating SARS-CoV-2-specific T cells in infection and vaccination. Unfortunately, none of the combination of antigens provided by the manufacturer enable identification of previous infection in vaccinated individuals, because all three antigen sets contain spike peptides.

There was little evidence to support a strong correlation between the QuantiFERON SARS-CoV-2 assay tubes and the ELISpot assay. Interestingly, there was statistically significant correlations between QuantiFERON SARS-CoV-2 and the MSD antibody data, however R squared values were relatively low, therefore strong conclusions about a true relationship between the sample sets cannot be drawn but may merit further investigation with a larger sample set.

The primary aim of this study was to determine the utility of the commercially-available QuantiFERON SARS-CoV-2 assay in evaluating T cells following SARS-CoV-2 infection and vaccination. The data demonstrates this assay, particularly the Ag3 assay tube, to be highly sensitive and specific in detecting SARS-CoV-2 T cells in acute but not past infection, and capable of discerning differences in T cell responses in unvaccinated and vaccinated individuals albeit with reduced sensitivity compared to the PITCH ELISpot. The assay was also relatively easy to perform, using equipment commonly available in a hospital laboratory. Therefore, the assay may be beneficial in laboratories which do not have access to established T cell assays, and as a dichotomous measure for monitoring of vaccine immunogenicity. Further research is required to define a suitable timeline following infection or vaccination during which the QuantiFERON SARS-CoV-2 assay may be applied to detect SARS-CoV-2-specific T cells as well as further development of the platform to increase the sensitivity for SARS-CoV-2 T cell responses in more distant infection and vaccination.

## Limitations

This study has a number of limitations. Firstly, genotype data for past infections was unavailable, although we know that the previously infected participants were chiefly infected by early pandemic strain virus in wave 1, and the acute infection group were infected when the delta variant was predominant (32). Further testing in populations with documented different variants is required, although T cell responses have been shown to be only marginally impacted by alpha, beta, gamma and delta variants (5,6) and 75-85% preserved in Omicron (44–50). This study involved only a single time point, without samples between the acute and previous infection timepoints. Furthermore, for naïve samples, we cannot rule out the possibility that asymptomatic infection may have occurred previously as antibodies reduce significantly over time (8). This study enrolled young (ages 18-56) and healthy individuals, whilst T cell response to vaccination is known to be affected by aging (51) and immunosuppression (52,53). This study was biased towards female participants, although larger studies have not found sex to be a determinant of SARS-CoV-2-specifiic T cell responses (5). There was also limited ethnic diversity in this cohort. Finally, the number of participants recruited may have rendered some of the statistical analysis sub-optimal, particularly in regard to correlation analysis with a significant proportion of zero values in the QuantiFERON assays. The limitations suggest further larger studies with genetically sequenced strains with a population to include a range of SARS-CoV-2 variants, ages, co-morbidities, sexes and ethnicities are warranted.

## Data Availability

The data underlying this article are available in the article and in its online supplementary material.

## Ethics Approval Statement

Participants were enrolled in the OPTIC study (GI Biobank Study 16/YH/0247, approved by the research ethics committee (REC) at Yorkshire & The Humber - Sheffield Research Ethics Committee on 29 July 2016, and amended for the OPTIC study on 8 June 2020 or the Family Study R71346/RE001, approved by Oxford University’s Medical Sciences Inter-Divisional REC (MS-IDREC-R71346/RE00).

## Patient Consent Statement

All participants in the study gave written, informed consent.

## Permission to Reproduce Material from Other Sources

Not applicable

## Clinical Trial Registration

Not applicable

## Conflict of Interest Disclosure

S.J.D declares fees as a Scientific Advisor to the Scottish Parliament on COVID-19. No other competing interests declared.

QuantiFERON assays were purchased as part of a commercial contract and the company played no role in this report.

## Acknowledgements

We are grateful to all our healthcare worker colleagues who participated in the study, and to Lisa Fielding for administrative support.

## Author Contributions

SAJ, SJD, EB and PK conceptualised the study. SAJ, SJD, EB, MC, KJ, CPC and PK designed and oversaw the clinical study. SAJ, CM, LS, AJ, JW and SJD recruited participants and collected samples. SAJ, EP, SA, SL, TM, MA, DTS implemented laboratory testing. SAJ and JN undertook data analysis. AD undertook project administration. SJD, PK and EB acquired the funding. SAJ, JN, EJB, PK and SD prepared the manuscript, which was reviewed by all contributing authors.

---

**Supplementary Figure 1.**
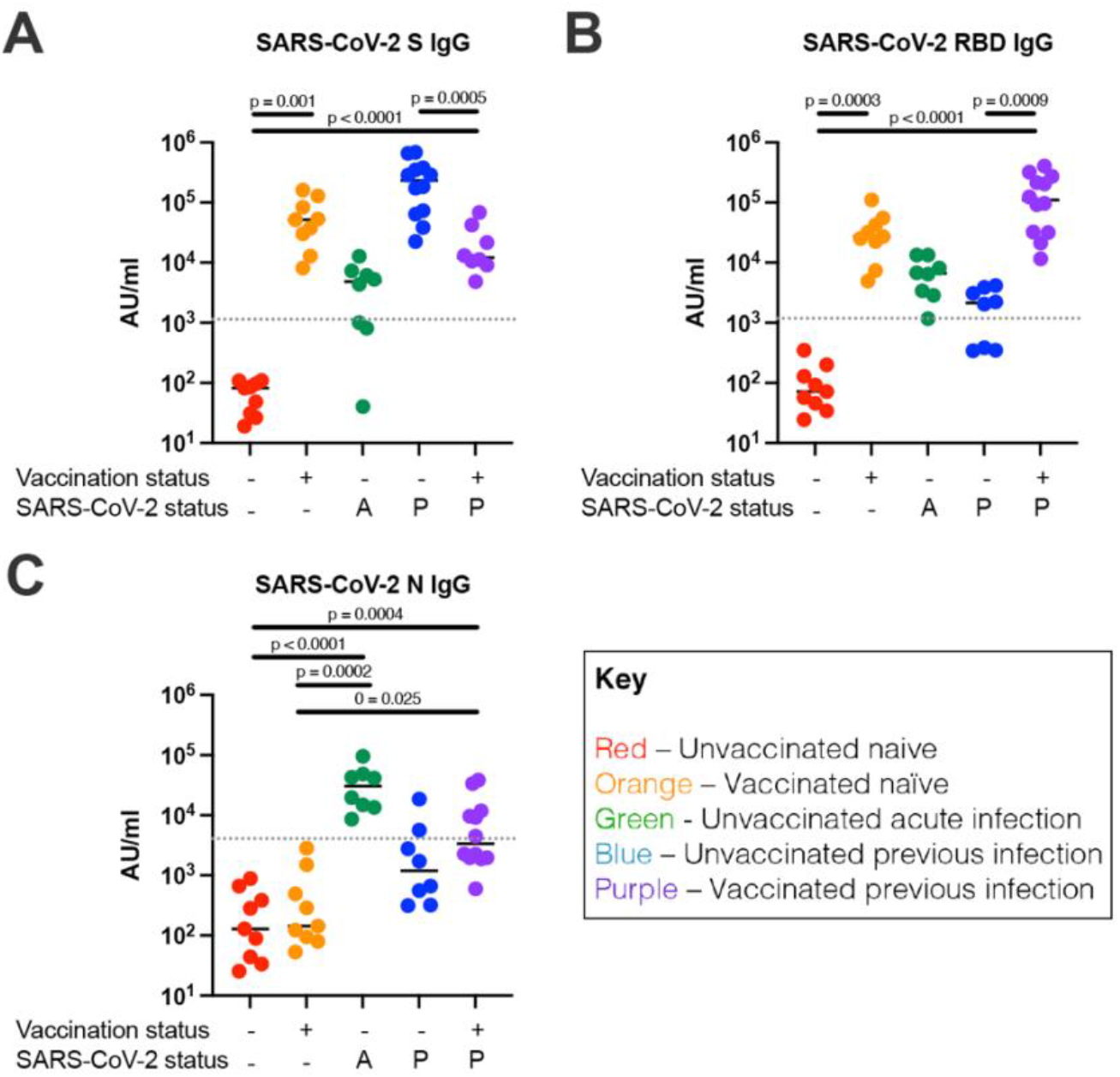
SARS-CoV-2 S-, RBD- and N-specific IgG responses using MSD binding assay. (A) S IgG antibody titres for individual groups. (B) RBD IgG antibody titres for individual groups. (C) N IgG antibody titres for individual groups. Unvaccinated naïve, n = 9; unvaccinated acute infection, n = 8; unvaccinated previous infection, n = 8; vaccinated naïve, n = 10; vaccinated previous infection, n = 12. A – acute infection; P – previous infection. Unpaired comparisons between groups were performed using Kruskal-Wallis test with Dunn’s correction. Horizontal dotted lines represent the threshold of each assay based on pre-pandemic controls (see Methods).

**Supplementary Figure 2.**
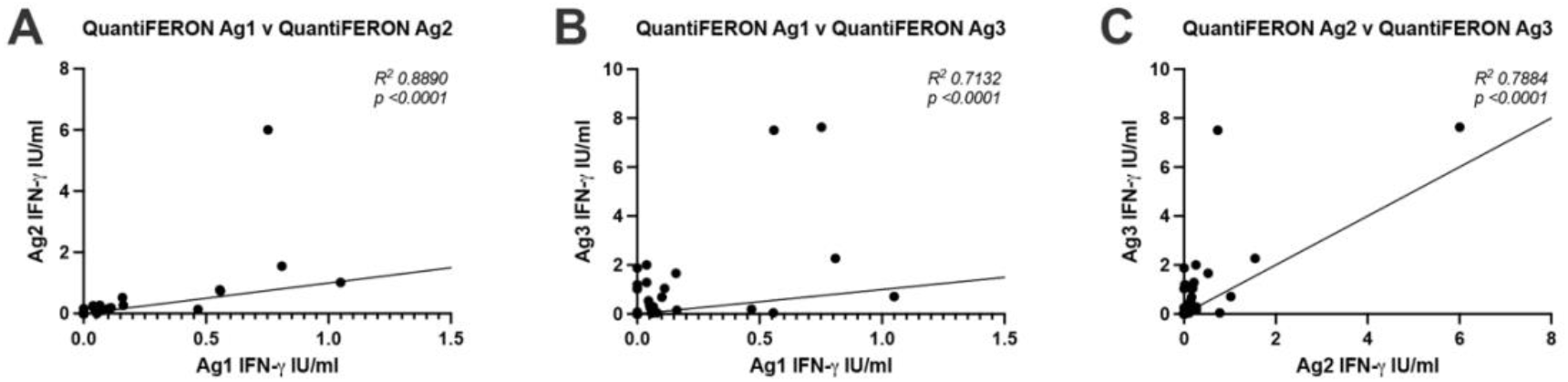
Correlation between T cell responses measured by QuantiFERON assay tubes. (A) Correlation between Ag 1 v Ag2 (B), Ag1 v Ag3 and (C) Ag2 v Ag3. N = 47 for each analysis. Correlation analysis was carried out with Spearman’s r correlation and two-tailed P values reported, with alpha = 0.05.

**Supplementary Figure 3.**
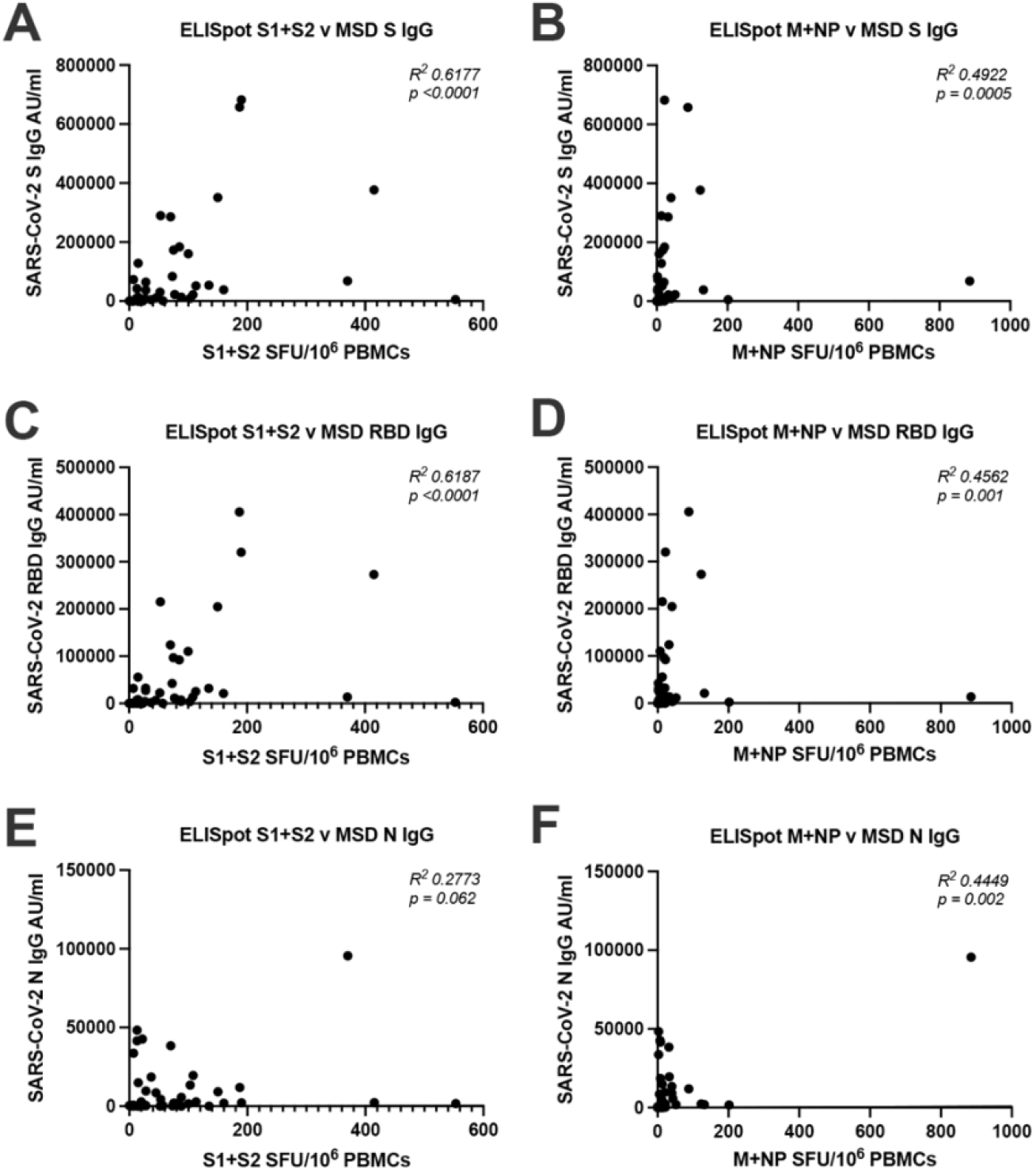
Correlation between T cell responses measured by ELISpot and antibodies measured by MSD binding assay. (A) Correlation analysis between ELISpot S1+S2 and MSD S IgG (B) ELISpot M+NP and MSD S IgG (C) ELISpot S1+S2 and MSD RBD IgG (D) ELISpot M+NP and MSD RBD IgG (E) ELISpot S1+S2 and MSD N IgG (F) ELISpot M+NP and MSD N IgG. N = 47 for each analysis. Correlation analysis was carried out with Spearman’s r correlation and two-tailed P values reported, with alpha = 0.05.

**Supplementary Figure 4.**
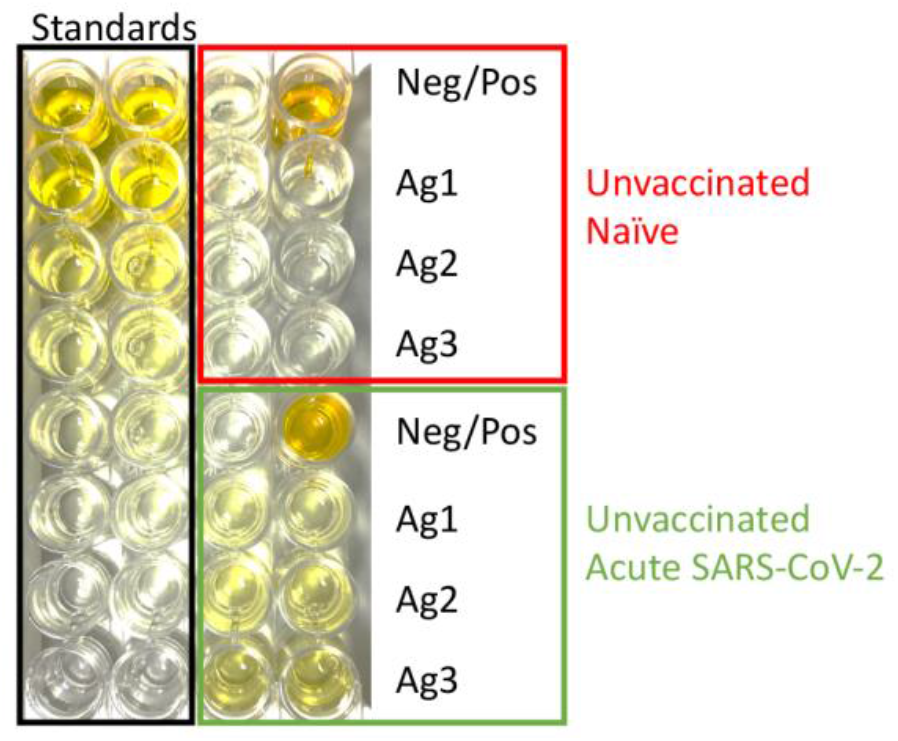
Visual representation of the QuantiFERON IFN-γ ELISA plate. Standards are depicted in duplicate in the first two columns (black box). Wells from a sample of an unvaccinated, naïve participant are depicted (red box) with a negative control (top left), positive control (top right) and the experimental wells of Ag1, Ag2 and Ag3 in duplicate. Wells from a sample of an unvaccinated, acute infection participant are depicted (green box) are similarly presented.

